# Using Machine Learning and Electronic Health Record (EHR) Data for the Early Prediction of Alzheimer’s Disease and Related Dementias

**DOI:** 10.1101/2024.12.09.24318740

**Authors:** Sonia Akter, Zhandi Liu, Eduardo J. Simoes, Praveen Rao

**Affiliations:** Institute for Data Science and Informatics, University of Missouri, USA; Department of Electrical Engineering and Computer Science, University of Missouri, USA; Department of Biomedical Informatics, Biostatics, and Medical Epidemiology, University of Missouri, USA

**Keywords:** Alzheimer’s Disease Dementias, Machine Learning (ML), Electronic Health Record data Early Prediction

## Abstract

**Objective:** **In the United States**, over 6 million patients are affected by Alzheimer’s Disease and Related Dementias (ADRD). The study aims to develop and validate machine learning (ML) models for the early diagnosis and prediction of ADRD using de-identified Electronic Health Record (EHR) data from the University of Missouri (MU) Healthcare for different prediction windows.

**Materials and Methods:** The study used de-identified EHR data provided by the MU NextGen Biomedical Informatics (BMI). An initial cohort of 380,269 patients aged over 40 with at least two healthcare encounters was narrowed to a final dataset of 4,012 unique patients of ADRD cases and 119,723 unique patients of controls. We trained and evaluated six different ML classifier models: Gradient-Boosted Trees (GBT), Light Gradient-Boosting Machine (LightGBM), Random Forest (RF), eXtreme Gradient-Boosting (XGBoost), Logistic Regression (LR), and Adaptive Boosting (AdaBoost) using metrics such as Area Under the Receiver Operating Characteristic Curve (AUC-ROC) score, accuracy, sensitivity, specificity, and F1 score. SHAP (SHapley Additive exPlanations) analysis was used to interpret predictions.

**Results:** The GBT model achieved the best AUC-ROC scores of 0.809, 0.821, 0.822, 0.808, and 0.833 for 1-year, 2-year, 3-year, 4-year, and 5-year prediction windows, respectively. The SHAP analysis highlighted key risk factors for ADRD, including depressive disorder, heart disease, higher age, headache, anxiety, and insomnia.

**Conclusion:** This study demonstrates the potential of ML models using EHR data for early ADRD prediction, enabling timely interventions to delay progression and improve outcomes. These findings offer insights for future research and proactive care strategies.

## INTRODUCTION

Alzheimer’s Disease and Related Dementias (ADRD) are a group of irreversible neurodegenerative disorders that progressively impair cognitive functions, memory, and the ability to perform daily activities^1,2^. Alzheimer’s Disease (AD) was first described by a German psychiatrist Alois Alzheimer. It was observed in a patient called Auguste who died in 1906 due to the loss of cognitive function^3^. Biologically, AD is defined as the pathological deposition of amyloid-beta (Aβ), tau proteins, and neurodegeneration in the brain^4–7^. These pathological changes often emerge 20 years before clinical symptoms appear^8^. As the disease advances, it progresses from Mild Cognitive Impairment (MCI) to severe dementia, with limited treatment options^9–11^.

In 2022, over 6 million Americans aged 65 or older were living with AD, and it was the seventh leading cause of death^12^. ADRD imposes a significant burden on patients, families, and society^13,14^. By 2030, the number of AD patients is expected to exceed to 75 million and double by 2050^12,13,15^. In 2022, the treatment for AD and dementia cost $321 billion, along with an additional $271 billion in unpaid caregiving, with projected annual costs exceeding $1 trillion by 2050^13,16^.

Early detection of ADRD is essential, as it allows for intervention before major cognitive decline takes place. Despite extensive research efforts, nearly 99% of clinical trials failed between 2002 and 2012 to develop successful treatments for ADRD^17^. However, diagnosis at the mild cognitive impairment (MCI) stage instead of late dementia could save the U.S. Healthcare System up to $7.9 trillion^12,18,19^. Since 1998, the U.S. Food and Drug Administration (FDA) has approved only six drugs to relieve ADRD symptoms: Rivastigmine, Galantamine, Donepezil, Memantine, Aducanumab, and a combination of Memantine and Donepezil^20–22^. These FDA-approved medications are most effective in the early to middle stages of the disease, offering symptomatic relief but not halting disease progression^17,20–23^. The limited success of these treatments highlights the need for more effective therapeutic strategies.

The emergence of machine learning (ML) offers new hope for improving the early detection of ADRD. By leveraging vast datasets from electronic health record (EHR) systems, administrative claims, and neuroimaging, ML models can uncover patterns and insights that may not be apparent through traditional methods. Recent studies have demonstrated the potential of ML in predicting AD incidence and managing other neurological conditions^24–28^. Previous studies have demonstrated the significant adaptability of ML models for different datasets, highlighting their robustness and applicability to other critical conditions such as cardiovascular disease, breast cancer, and prostate cancer^29–31^.

In our study, we applied six different ML models including Gradient-Boosted Trees (GBT)^32^, Light Gradient-Boosting Machine (LightGBM)^33^, Random Forest (RF), eXtreme Gradient-Boosting (XGBoost)^34^, Logistic Regression (LR)^35^, and Adaptive Boosting (AdaBoost)^36^ to classify ADRD using de-identified EHR data from the University of Missouri (MU) Healthcare, which is part of the National Patient-Centered Clinical Research Network (PCORnet). By optimizing the ML models for accuracy, Area Under the Receiver Operating Characteristic Curve (AUC-ROC), sensitivity, and F1scores, and by identifying key predictive features, we aimed for accurate and early diagnosis of ADRD. We employed all captured EHR variables as predictors and tested different prediction windows spanning 1 to 5 years. The key findings of this study are as follows: The GBT model outperformed other classifiers across multiple prediction windows. SHAP (SHapley Additive exPlanations) analyses highlighted important risk factors for ADRD, which included depressive disorder, age, heart disease, anxiety, headache, and sleep apnea. The findings from this study have the potential to advance diagnostic tools, supporting clinicians in making more precise decisions and improving outcomes for patients with ADRD.

In the remaining section of the paper, we will delve into the methodology, and the results, and discuss the significance of the findings.

## METHODS

We used de-identified EHR data from MU Healthcare, provided by the MU NextGen BMI. The data were modeled using the PCORnet Common Data Model (CDM)^37^, containing longitudinal EHRs encompassing diverse patient characteristics, including demographics, diagnoses, medications, vital signs, and smoking history. This study was approved by the MU Institutional Review Board under protocol IRB2095682.

### Study Participants

This retrospective case-control study focused on predicting ADRD diagnosis in adults aged 50 years and older. We began with a cohort of 380,269 patients who met the following criteria: (1) aged 40 years or older as of January 1, 2010 (the start date of records in the MU EHR system), (2) admitted between January 1, 2010, and December 31, 2023, and (3) had at least two recorded encounters in MU Healthcare.

### Selection Criteria for Cases and Controls

In this study, we defined ADRD cases using two main criteria for prediction. First, patients who had an ADRD diagnosis based on the International Classification of Diseases, 9th or 10th Revisions (ICD-9/ICD-10), which included codes 331.0, 290.0, 290.1, 290.2, 290.3, 290.4, 290.43, 331.82, 294.1, G30.0, G30.1, G30.8, G31.83, F00, F00.2, F01, F02, and F00.9. These ICD codes cover early onset, late onset, and confirmed ADRD cases (that do not specifically fit into early or late onset categories). We also included patients who were prescribed dementia-related medications commonly used for AD, such as rivastigmine, galantamine, donepezil, memantine, brexpiprazole, and aducanumab. Second, patients must have recorded at least two encounters in the MU EHR system.

For the control group (non-ADRD), we selected individuals who (1) had no ADRD-related diagnoses based on ICD-9/10 codes or were not prescribed any dementia-related medications, and (2) had at least two recorded encounters in the MU EHR system. Our final dataset comprised of 123,735 unique patients; of these, 4,012 were diagnosed with ADRD (cases), while 119,723 had no ADRD or ADRD-related diagnoses (controls).

### Study Design

In our study, we divided the time into two sections: (1) an observation window and (2) a prediction window. We set an index date for each case as the earliest date of either an ADRD diagnosis or the first prescription of a dementia-related medication. We defined multiple prediction windows: 1 year, 2 years, 3 years, 4 years, and 5 years, representing the start of a recorded ADRD diagnosis or the first prescription of dementia-related medication. The observation phase spanned from January 1, 2010 (the start date of records in the MU EHR system) to December 31, 2018. Data from the observation phase were exclusively used for prediction, allowing us to evaluate the potential for forecasting ADRD at different time intervals.

We used a fixed control dataset (2010-2018) and different case datasets for each prediction window as shown in Figure 1. The fixed control dataset was the historical data from which the model learned, remaining consistent across all prediction windows. This approach provided a stable base for our model to understand patterns and trends up to 2018, allowing us to isolate the effect of varying prediction windows (case datasets) on model performance. For instance, if a patient was diagnosed with ADRD in 2019, the control dataset would remain the same (2010-2018), while the case dataset would adjust according to the prediction window (e.g., 1-year prediction: 2019-2020, 2-year prediction: 2019-2021, and so on). To address the presence of multiple test measurements from different encounters, the most recent measurements before the end of the observation period were selected for analysis.

**Figure 1.**
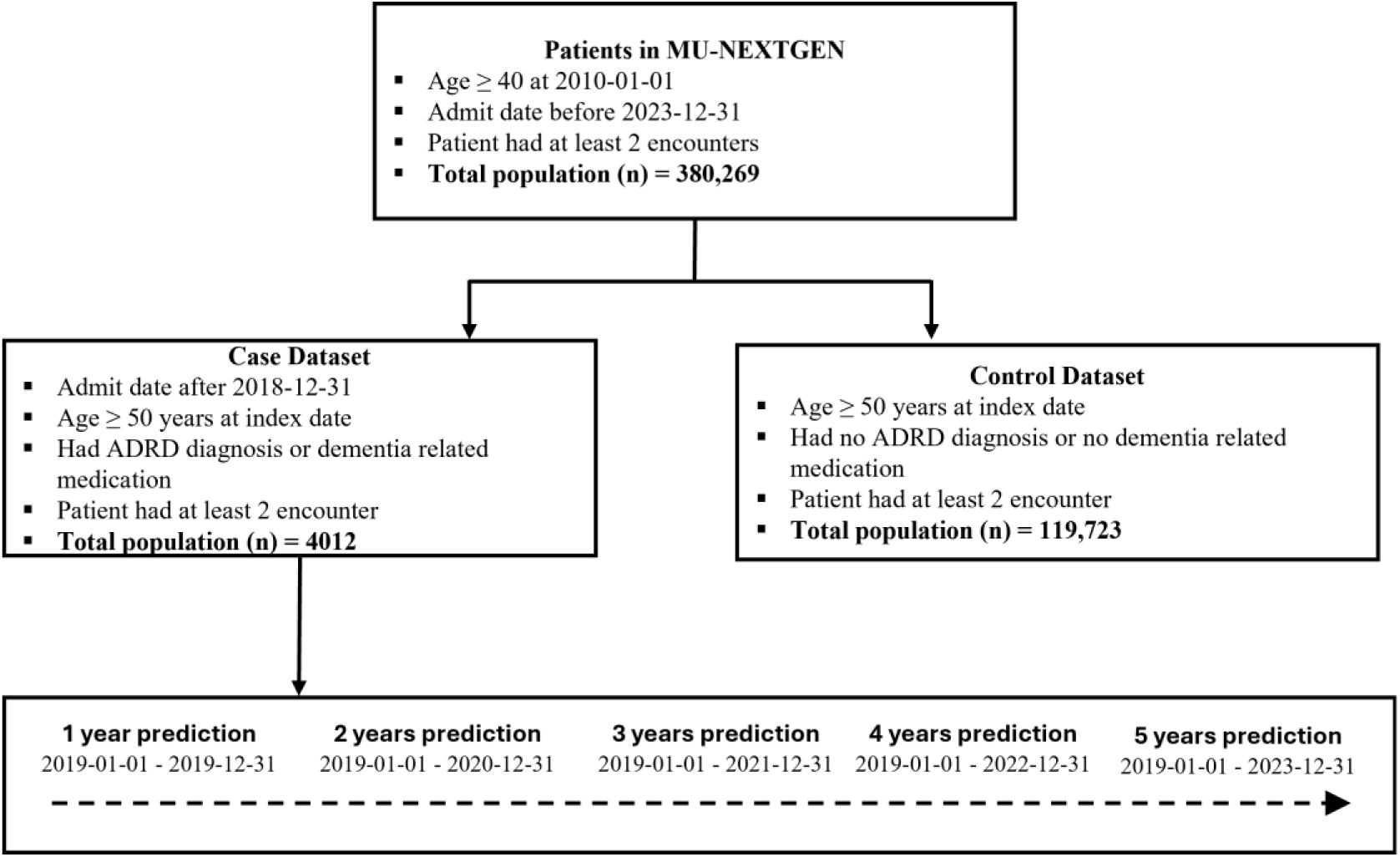
Flowchart for preparing the case-control study.

Our dataset included variables captured by the EHR system, such as (1) demographic variables (age, race, marital status, sex, and smoking history) and (2) vital variables, namely, diastolic blood pressure (DBP) and systolic blood pressure (SBP). Additionally, we incorporated comorbidities as risk factors, identified through a thorough review of existing literature^23,38^. The comorbidities and medical diagnoses include diabetes, epilepsy, depression, obesity, stroke, anxiety, hypertension, hyperlipidemia, cardiovascular disease, sleep disorder, headache, periodontitis, concussion, heart disease, sleep apnea, insomnia, kidney disease, cholesterol, vitamin D deficiency, enlarged prostate, bone disease, and depressive disorder.

### Data Pre-processing

To address missing data, we excluded the feature variables with a missing rate of 30% or higher. Patients with more than 20% missing data were removed from the analysis, while those with less than 20% missing data were imputed. Initially, our dataset included 4,012 unique patients in the positive class (cases), and 232,795 unique patients in the negative class (controls), the final number of cases and controls is shown in Figure 1.

In the data preprocessing phase, one-hot encoding was applied to all categorical variables. Continuous variables were handled according to their specific characteristics. For example, Age was categorized into five distinct categories: [50,60), [60,70), [70,80), [80,90), and above 90 years. DBP and SBP were categorized into three levels based on clinical thresholds: ‘normal,’ ‘high,’ and ‘critically high.’ Specifically, DBP was categorized as normal (<80 mm Hg), high (80-90 mm Hg), and critically high (>90 mm Hg), while SBP was categorized as normal (<120 mm Hg), high (120-140 mm Hg), and critically high (>140 mm Hg)^39^.

The resulting feature vector consisted of binary values, where 0s and 1s indicated the absence or presence of each category. For medical diagnosis or comorbidities conditions, one-hot encoding was used to construct the feature vector. The smoking history was encoded into binary values: ‘never smoker’ was mapped to 0, while all other smoking categories, including former and current smokers, were mapped to 1. This approach enabled a simplified distinction between non-smokers and those with any smoking history.

The dataset was divided into training and testing sets, with 80% for training and the remaining 20% for testing. The training set was used to develop the models, while the testing set was employed to assess their performance.

### Model Validation and Analysis

We trained and tested six different ML classification models: GBT, LightGBM, RF, XGBoost, LR, and AdaBoost to predict ADRD at an early stage. A nested cross-validation approach was employed to optimize and evaluate these models. Each model was incorporated into a pipeline that included a StandardScaler for feature normalization, followed by the respective classifier.

Hyperparameter tuning was conducted using a 5-fold StratifiedKFold inner cross-validation loop with grid search (GridSearchCV). The optimal hyperparameters were then applied in an outer 5-fold StratifiedKFold cross-validation to assess model performance. Predicted probabilities were classified using a 0.5 threshold, and the model performance was measured using metrics such as accuracy, precision, sensitivity, F1 score, AUC-ROC, and specificity, with confusion matrices generated for each fold. Bootstrapping with 1,000 iterations was applied to estimate point values. The model with the best performance across all metrics was selected and further evaluated on a hold-out test set to assess generalization.

We applied SHAP (Shapley Additive exPlanations) methods^40^ to interpret the predictions of the ML models, we generated SHAP values for each of the five prediction windows (1, 2, 3, 4, and 5 years). These values were used to generate summary plots, providing insights into the model interpretability and highlighting risk factors over time. Specifically, SHAP bar plots and summary plots were generated for the top 12 risk factors in the best-performing model. Features with positive SHAP values were linked to a higher probability of ADRD, whereas those with negative values were associated with a lower risk. The magnitude of each SHAP value reflected the overall importance of that feature with the model.

## RESULTS

### Sample Characteristics

Table 1 shows the descriptive statistics of the case and control groups. Our dataset includes 119,723 control individuals and 4,012 cases, all aged 50 years and older, covering the period from January 1, 2010, to December 31, 2023. The analysis focused on key demographic characteristics of both groups. The mean age in the case group (77.50 ± 9.25 years) was higher than in the control group (75.51 ± 10.18 years), indicating an older population in the case group. Female patients were more prevalent than male patients in both groups. Additionally, the White race was predominant in both groups given the demographics of patients visiting MU Healthcare.

**Table 1.**
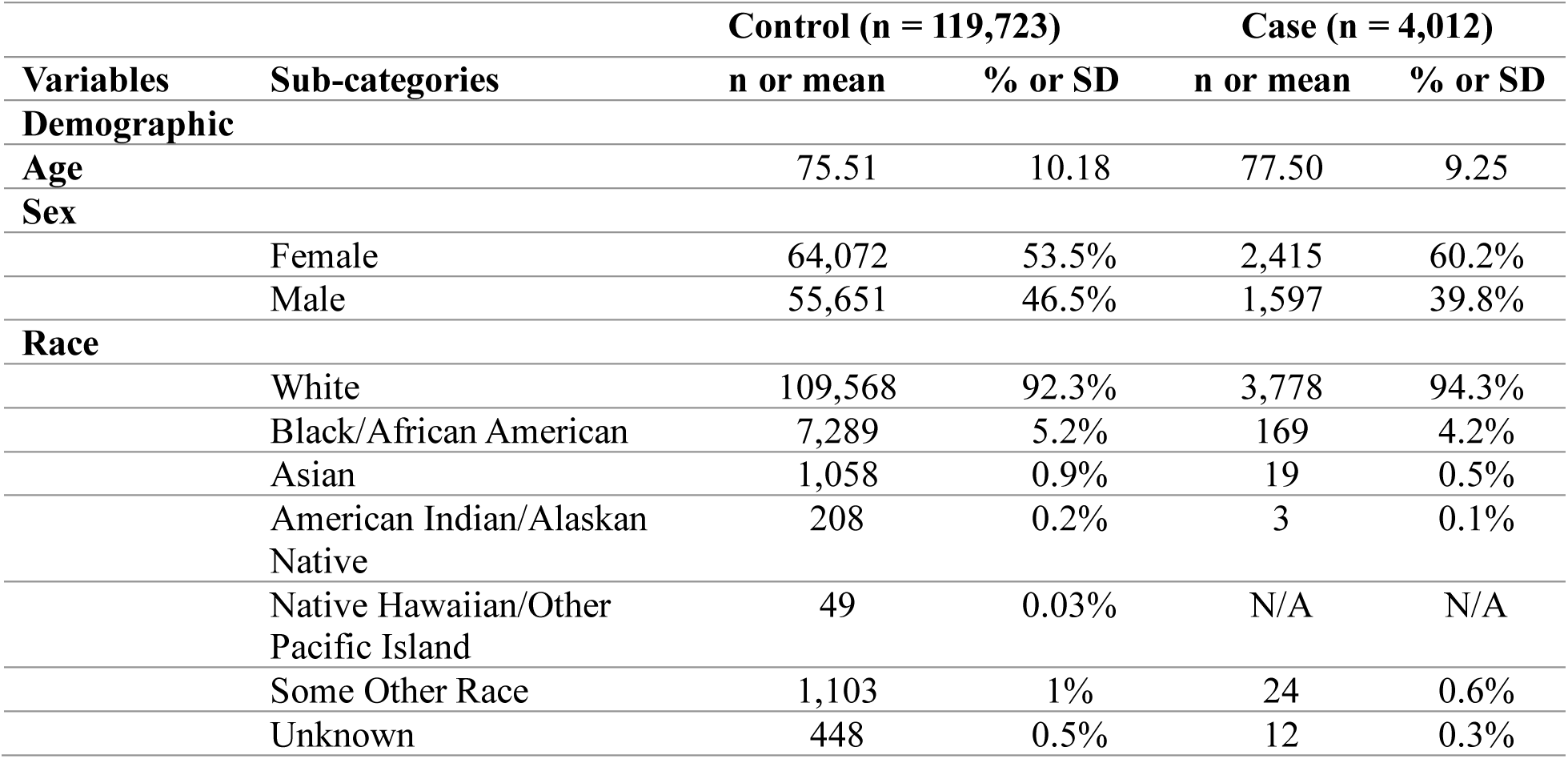
Descriptive Statistics in Control and Case.

### Performance Evaluation of Model Prediction

We trained six different ML classification models namely: LR, GBT, LightGBM, XGBoost, RF, and AdaBoost by applying hyperparameter tuning on the training set. These models were trained to predict ADRD incidence over 1-year, 2-year, 3-year, 4-year, and 5-year prediction windows. Using the best performing model for each classifier, we classified the unseen test set into classes: ADRD (case, positive class) and non-ADRD (control, negative class) patients.

Next, we report the ML model performance. As shown in Table 2, the Area under the curve (AUC) using the GBT model demonstrated the highest performance in predicting ADRD over 5 years. The GBT model consistently outperformed the other model in terms of AUC and accuracy in all 5 prediction windows. The GBT model achieved the best AUC score of 0.833, 808, 0.822, 0.821, and 0.809 for predicting ADRD at 5-year, 4-year, 3-year, 2-year, and 1-year prediction windows, respectively. LightGBM and XGBoost also exhibited strong performance, with AUC scores of 0.831 and 0.829, respectively in the 5-year prediction window. In contrast, the LR model had the lowest AUC of 0.782 in the 5-year prediction window. The AUC score consistently increased as the prediction window extended from 1-year to 5-year. Similar results were obtained for other performance metrics, including accuracy, sensitivity, specificity, and F1 scores.

**Table 2.** Performance of ADRD Predictive Models (^a^ best model according to AUC score).

| Prediction Window | Model | Accuracy | AUC | Precision | Sensitivity | Specificity | F-1 |
| --- | --- | --- | --- | --- | --- | --- | --- |
| 1 Year | LR | 0.696 | 0.775 | 0.970 | 0.696 | 0.695 | 0.801 |
|  | GBT | 0.978 | 0.809<sup>a</sup> | 0.970 | 0.978 | 0.999 | 0.968 |
|  | LightGBM | 0.970 | 0.808 | 0.964 | 0.970 | 0.999 | 0.957 |
|  | XGBoost | 0.979 | 0.799 | 0.975 | 0.979 | 1.000 | 0.970 |
|  | RF | 0.978 | 0.792 | 0.978 | 0.978 | 1.000 | 0.967 |
|  | AdaBoost | 0.978 | 0.782 | 0.969 | 0.978 | 1.000 | 0.968 |
| 2 Years | LR | 0.684 | 0.787 | 0.965 | 0.684 | 0.682 | 0.789 |
|  | GBT | 0.974 | 0.821<sup>a</sup> | 0.969 | 0.974 | 1.000 | 0.962 |
|  | LightGBM | 0.973 | 0.821 | 0.963 | 0.973 | 0.999 | 0.962 |
|  | XGBoost | 0.976 | 0.818 | 0.972 | 0.976 | 0.999 | 0.967 |
|  | RF | 0.974 | 0.816 | 0.974 | 0.974 | 1.000 | 0.961 |
|  | AdaBoost | 0.974 | 0.796 | 0.964 | 0.974 | 0.999 | 0.962 |
| 3 Years | LR | 0.688 | 0.784 | 0.961 | 0.688 | 0.687 | 0.789 |
|  | GBT | 0.974 | 0.822<sup>a</sup> | 0.971 | 0.971 | 0.999 | 0.965 |
|  | LightGBM | 0.971 | 0.817 | 0.964 | 0.971 | 0.999 | 0.958 |
|  | XGBoost | 0.974 | 0.798 | 0.970 | 0.974 | 0.998 | 0.966 |
|  | RF | 0.971 | 0.807 | 0.972 | 0.971 | 1.000 | 0.958 |
|  | AdaBoost | 0.970 | 0.800 | 0.941 | 0.970 | 1.000 | 0.955 |
| 4 Years | LR | 0.698 | 0.763 | 0.956 | 0.698 | 0.698 | 0.795 |
|  | GBT | 0.970 | 0.808<sup>a</sup> | 0.964 | 0.970 | 0.999 | 0.957 |
|  | LightGBM | 0.970 | 0.808<sup>a</sup> | 0.964 | 0.970 | 0.999 | 0.957 |
|  | XGBoost | 0.971 | 0.807 | 0.966 | 0.971 | 0.999 | 0.960 |
|  | RF | 0.969 | 0.797 | 0.968 | 0.969 | 0.999 | 0.954 |
|  | AdaBoost | 0.969 | 0.795 | 0.960 | 0.969 | 0.999 | 0.956 |
| 5 Years | LR | 0.688 | 0.782 | 0.955 | 0.688 | 0.687 | 0.787 |
|  | GBT | 0.970 | 0.833<sup>a</sup> | 0.968 | 0.970 | 0.999 | 0.960 |
|  | LightGBM | 0.969 | 0.831 | 0.966 | 0.969 | 0.999 | 0.958 |
|  | XGBoost | 0.968 | 0.829 | 0.961 | 0.969 | 0.999 | 0.953 |
|  | RF | 0.968 | 0.823 | 0.967 | 0.968 | 0.999 | 0.954 |
|  | AdaBoost | 0.967 | 0.818 | 0.959 | 0.967 | 0.999 | 0.953 |

The AUC-ROC curves for the 5-year prediction window using six different ML models are displayed in Figure 2 (right). For example, the GBT model achieved an AUC score of 0.833, followed by the LightGBM, XGBoost, RF, AdaBoost, and LR with an AUC score of 0.831, 0829, 0.822, 0.818, and 0.782, respectively. Additionally, Figure 2 (left) displayed the AUC-ROC curves for predictions across 1-year, 2-year, 3-year, 4-year, and 5-year prediction windows using the best model, GBT. This upward trend from 1-year to 5-year suggests that the predictive accuracy of the GBT model helps with the inclusion of longitudinal data. The findings were consistent across other metrics, including specificity and sensitivity.

**Figure 2.**
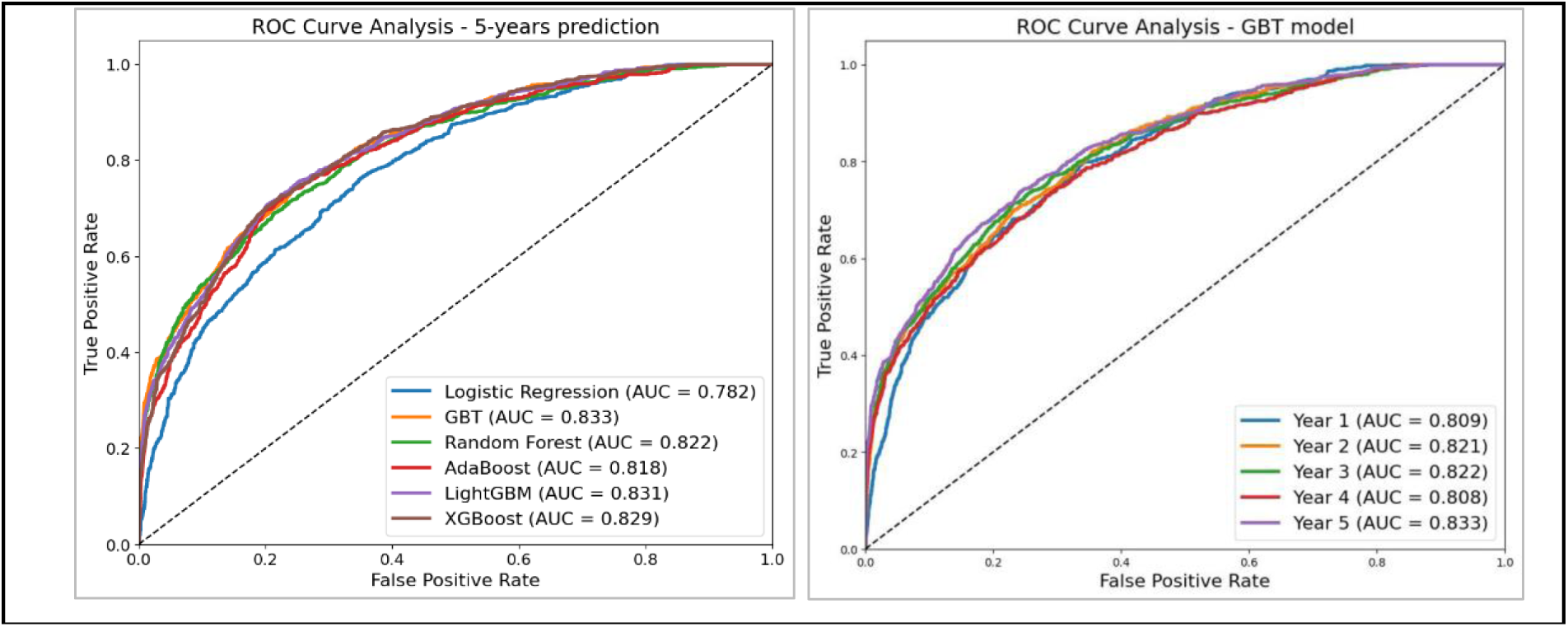
Performance assessment of ML models in ADRD prediction.

ROC curve analysis for the 5-year prediction window using six different ML models (Left), ROC curve analysis for predictions across 1-, 2-, 3-, 4-, and 5-year windows. The GBT model, being the best performer, was used for the ROC plot (Right).

### SHAP Analysis and Model Interpretability

We applied the SHAP to identify the key risk factors influencing ADRD prediction and their relationship with outcomes. Given the GBT model’s excellent performance, SHAP values provided insight into the model’s interpretability. Figure 3 presents the SHAP analysis across the 1-year to 5-year prediction window, identifying the top 12 features that most influenced the model’s predictions. Consistently, features such as a history of depressive disorder, higher age, history of anxiety, history of sleep apnea, history of heart disease, history of headache, and high DBP were the most significant predictors of ADRD risk.

**Figure 3.**
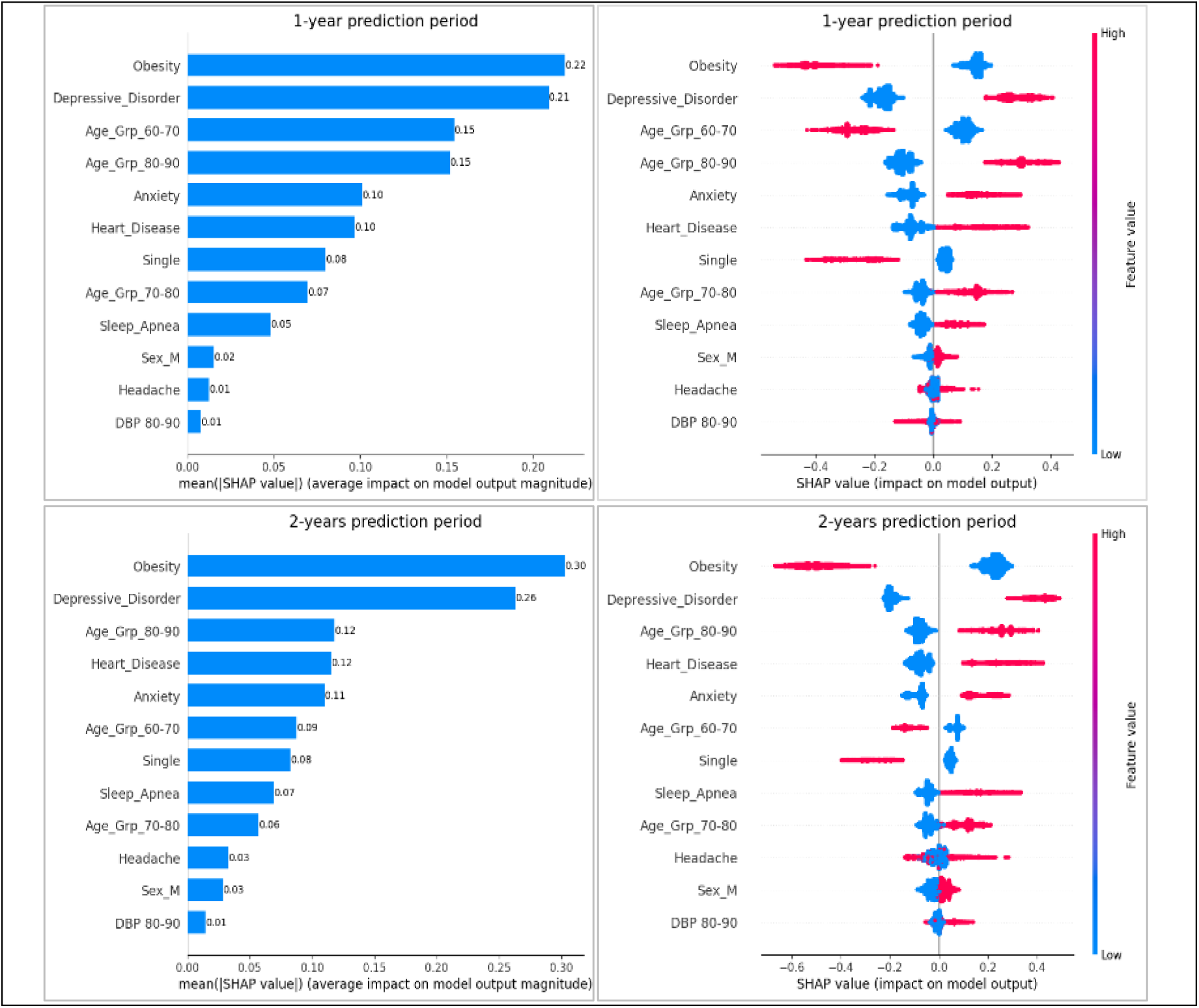
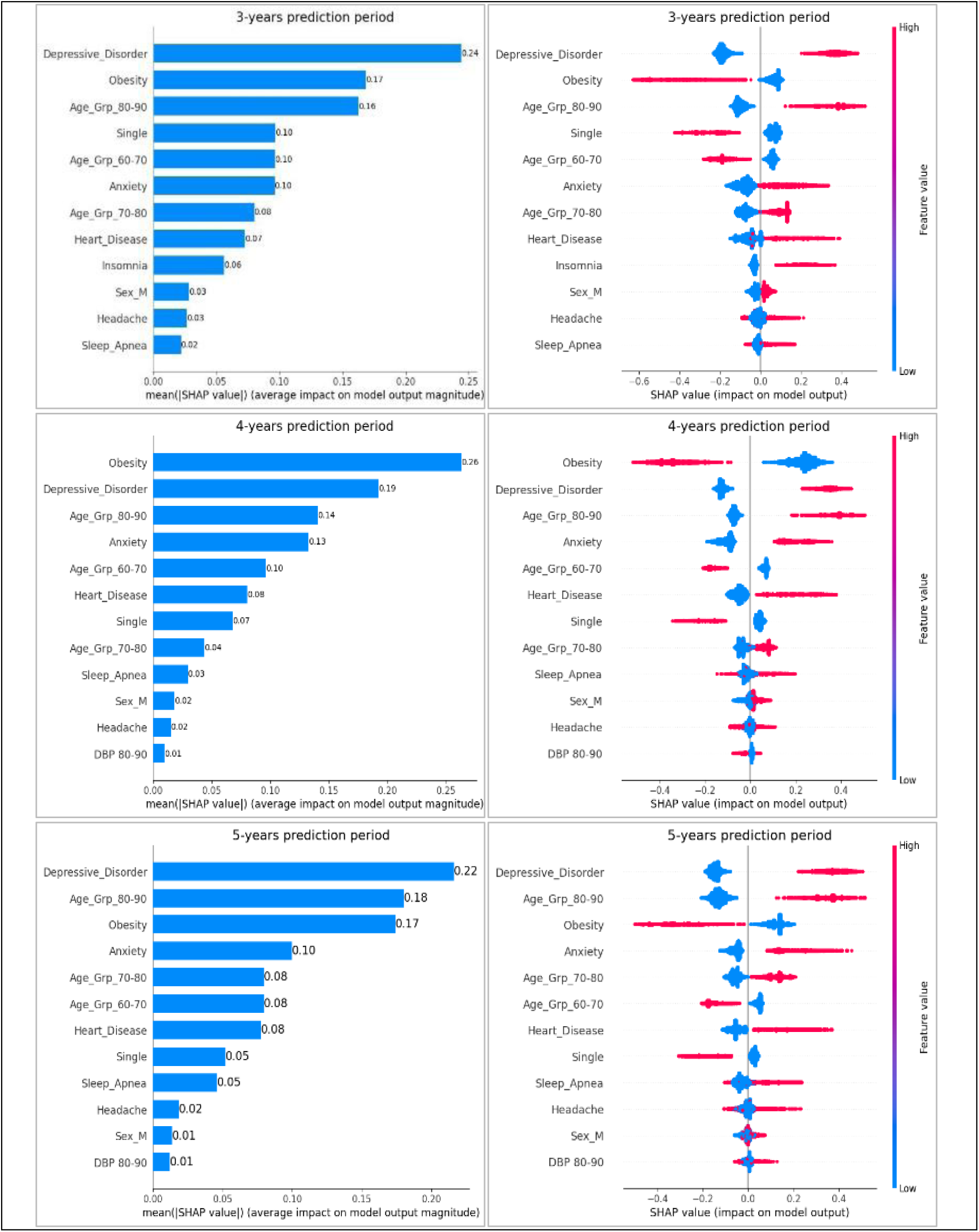
SHAP plots of the top-12 features for the GBT models (1-year - 5-year prediction windows).

The SHAP plot provides a detailed breakdown of how each feature affects the model’s prediction, highlighting the model’s interpretability and the complex relationships between different risk factors. Positive contributions, shown by the red segments, increased the likelihood of an ADRD prediction, while negative contributions, shown by the blue segments, decreased it.

## DISCUSSION

Our analysis of de-identified EHR data from MU Health Care, using ML models for early prediction of ADRD, revealed significant insights. We evaluated six models: GBT, LightGBM, RF, XGBoost, LR, and AdaBoost across different prediction windows: 1-year, 2-year, 3-year, 4-year, and 5-year before ADRD diagnosis. The GBT model, using features selected based on established literature consistently outperformed the others, achieving the highest AUC scores of 0.809, 0.821, 0.822, 0.808, and 0.833 for predictions made 1 to 5 years post-diagnosis, respectively. As the prediction window extended from 1 to 5 years, model performance generally improved, reflecting higher AUC scores. The 5-year prediction window demonstrated the best performance, capturing broader and more consistent trends, while the 1-year window, though effective, was more prone to noise and specific patterns that were harder to predict accurately.

In our study, the control dataset remains fixed (2010-2018) regardless of the prediction window, meaning that the amount of historical data used for training did not change. This contrasts with other studies ^23^, where the control dataset is expanded with more recent data as the prediction window shortens, likely making the predictions more accurate in the shorter timeframe. For example, their study stated that model performance declined as the prediction window increased from 0 to 5 years, with higher confidence in predicting ADRD onset closer to the initial diagnoses. This methodological difference in keeping a fixed dataset versus incorporating recent data could explain why our models performed better with a longer prediction window, as the broader 5-year window allowed for better generalization, leading to improved AUC performance.

By computing SHAP values using the best-performing model, we identified the most significant features contributing to ADRD prediction. Different ML models may utilize varying feature combinations due to distinct algorithmic assumptions, but our focus here is on the results from the top model. The SHAP analysis across multiple prediction windows (1-year, 2-year, 3-year, 4-year, and 5-year) provided critical insights into the model’s behavior and the relative importance of various risk factors, such as depressive disorder, age, obesity, anxiety, heart disease, relationship status (being single), headache, sleep apnea, high DBP, and insomnia. For example, depressive disorder was strongly associated with a higher risk of ADRD.

Notably, depressive disorder and heart disease emerged as significant risk factors, consistent with existing literature linking them to cognitive decline. Depression can negatively impact cognitive reserve, which is the brain’s ability to compensate for damage, while heart disease may reduce cognitive reserve through impaired blood flow and oxygenation. The bidirectional nature of these relationships is also important; cognitive decline in early ADRD may lead to depressive symptoms, which can further exacerbate dementia progression. Similarly, heart disease can contribute to depression, thereby increasing ADRD risk. Both conditions are associated with chronic inflammation and vascular damage, which play a role in ADRD development^41^.

Age, particularly in the 80-90 group, also significantly increased the predicted risk of ADRD, aligning with established research that highlights aging as a major non-modifiable risk factor for dementia. This association has been well documented in studies showing that dementia incidence continues to rise even in the oldest age group (≥ 90), where advanced age was found to be a significant non-modifiable risk factor for dementia^42^. Additional features such as anxiety, and sleep disorders also influenced the model’s predictions. The link between anxiety and increased ADRD risk supports research suggesting that chronic stress may accelerate cognitive decline, while the contribution of heart disease emphasizes the importance of vascular health in preserving cognitive function^43,44^.

In contrast to our study, previous research found indicators such as diabetes mellitus (both Type 1 and Type 2) to predict ADRD onset^45–47^. This difference may be due to variations in study populations, methodologies, or the specific features used in predictive models. Our findings suggest that other factors may play a more dominant role in ADRD risk within our cohort, highlighting the need for further investigation into the relationship between diabetes and ADRD across different populations.

Our SHAP analysis identified gender (male), headache, and insomnia as among the top 12 most significant risk factors for ADRD. This aligns with prior research linking headaches to increased dementia risk, suggesting a connection between chronic pain and cognitive decline^48^. Insomnia and other sleep disturbances, particularly among men, have also been associated with higher cognitive impairment risk^49,50^. These results underscore the importance of incorporating diverse risk factors, including those related to gender, sleep, and chronic pain, into ADRD predictive models. However, Li et al. did not find these factors significant in their study, emphasizing the variability in risk factors across different studies and the need to consider a broad range of variables in ADRD prediction models^23^.

Early identification of ADRD in clinical practice empowers healthcare providers, patients, and caregivers to take proactive measures. Healthy lifestyle changes, such as physical exercise, weight management, avoiding alcohol and sedatives, taking vitamins, and maintaining a healthy diet, can help control blood pressure and improve sleep, potentially reducing future ADRD risk. These findings highlight the multifactorial nature of ADRD risk, where psychological, metabolic, and demographic factors interact to influence disease onset. The SHAP analysis provides valuable insights into the model’s decision-making process, helping to identify the most predictive factors for ADRD at different timeframes. However, the unexpected relationship between obesity and ADRD risk warrants further investigation, as it challenges conventional assumptions and suggests a more complex interplay between metabolic health and cognitive decline than previously understood.

Identifying potential ADRD cases early through automated prediction models is crucial for timely referral to specialized care and guiding patient management. An effective approach may involve using a combination of models tailored to different stages of the disease. For instance, a long-term model could predict ADRD risk years in advance, while a short-term model could be employed as the disease progresses. These models not only give patients more time to prepare but also assist researchers in selecting and characterizing participants for clinical trials.

## CONCLUSION

In this study, we explored the use of ML to predict ADRD in patients using EHR data. We evaluated several well-known ML models, focusing on metrics such as accuracy, AUC-ROC, precision, and sensitivity. The GBT model outperformed other classifiers across multiple prediction windows. SHAP plot analyses revealed key risk factors, including depressive disorder, higher age, heart disease, anxiety, headache, and insomnia that are associated with ADRD risk. These findings highlight the potential of ML models to aid clinicians in identifying high-risk patients earlier, enabling proactive and targeted interventions that could improve patient outcomes and quality of life.

By facilitating early ADRD detection, our approach could help optimize treatment strategies, reduce healthcare costs, and enhance the support provided to both patients and caregivers. In the future, we aim to conduct a multi-center study to train ML models on a more diverse population, further improving their clinical applicability and generalizability.

## Acknowledgments

We thank MU NextGen Biomedical Informatics (BMI) for providing the data for this study.

## Authors’ contributions

SA, ZL, and PR designed the study. SA and ZL trained and evaluated ML models. SA, ZL, and PR analyzed the results. SA drafted the initial manuscript. SA, ZL, and PR edited the manuscript. ES provided feedback.

## Data availability statement

We cannot share the raw patient data. The NextGen BMI is responsible for the original data and the MU Hospital does not allow for sharing of the data because of HIPAA regulations.

